# Stress, Genetics and Mood: Impact of COVID-19 on a College Freshman Sample

**DOI:** 10.1101/2022.12.13.22283409

**Authors:** Cortney A Turner, Huzefa Khalil, Virginia Murphy-Weinberg, Megan H Hagenauer, Linda Gates, Yu Tang, Lauren Weinberg, Robert Grysko, Leonor Floran-Garduno, Thomas Dokas, Catherine Samaniego, Zhuo Zhao, Yu Fang, ijan Sen, Juan F Lopez, Stanley J Watson, Huda Akil

**Affiliations:** Michigan Neuroscience Institute, University of Michigan, Ann Arbor, MI, USA; Department of Psychiatry, University of Michigan, Ann Arbor, MI, USA; Department of Psychology, University of Michigan, Ann Arbor, MI, USA

**Keywords:** depression, students, polygenic risk

## Abstract

Using a longitudinal approach, we sought to define the interplay between genetic and non-genetic factors in shaping vulnerability or resilience to COVID-19 pandemic stress, as indexed by the emergence of symptoms of depression and/or anxiety. University of Michigan freshmen were characterized at baseline using multiple psychological instruments. Subjects were genotyped and a polygenic risk score for depression (MDD-PRS) was calculated. Daily physical activity and sleep were captured. Subjects were sampled at multiple time points throughout the freshman year on clinical rating scales, including GAD-7 and PHQ-9 for anxiety and depression, respectively. Two cohorts (2019-2021) were compared to a pre-COVID-19 cohort to assess the impact of the pandemic. Across cohorts, 26%-40% of freshmen developed symptoms of anxiety or depression (N=331). Depression symptoms significantly increased in the pandemic years, especially in females. Physical activity was reduced and sleep was increased by the pandemic, and this correlated with the emergence of mood symptoms. While Low MDD-PRS predicted lower risk for depression during a typical freshman year, this apparent genetic advantage was no longer evident during the pandemic. Indeed, females with lower genetic risk accounted for the majority of the pandemic-induced rise in depression. We developed a model that explained approximately half of the variance in follow-up depression scores based on psychological trait and state characteristics at baseline and contributed to resilience in genetically vulnerable subjects. We discuss the concept of multiple types of resilience, and the interplay between genetic, sex and psychological factors in shaping the affective response to different types of stressors.

**Significance Statement:** Biological and psychological factors that propelled the great rise in mood disorders during the COVID-19 pandemic remain unknown. We used a longitudinal design in three cohorts of college freshmen to parse the variables that contributed to susceptibility vs. resilience to pandemic stress. Low genetic risk (based on a depression polygenic risk score) was protective prior to the pandemic but this “genetic resilience” lost its effectiveness during the pandemic. Paradoxically, female students with low genetic risk showed enhanced vulnerability to depression during the pandemic across two cohorts. By contrast, we defined a baseline Affect Score (AS) comprising psychological variables that were predictive of future stress susceptibility or “psychological resilience” to stress even in the genetically vulnerable subjects.

## INTRODUCTION

Few longitudinal studies exist in a young population that define key determinants of stress vulnerability or resilience and capture the psychobiological characteristics of individuals before they transition to depression. Major Depressive Disorder (MDD) is highly genetically complex, with heritability estimates at 30-40% (1), indicating a strong role of environmental factors, with stress or life events often triggering the initial depressive episode (2, 3). Moreover, previous depressive episodes contribute to the likelihood of subsequent depression (4), which makes it more critical to predict and attempt to prevent the emergence of depression in young healthy individuals.

Parsing the relative role of genetic and psychosocial variables in the risk for depression remains a major challenge. Several studies have described a polygenic risk score for depression (MDD-PRS) (5), and a few have related this score to clinical outcomes (6–8). In youths, the MDD-PRS predicted depression severity and age of onset (7), consistent with Wray et al. (9). A recent study applied the MDD-PRS to medical interns (10), and pointed to internship stress as mediating the relationship between genetic risk and depression.

The current study focuses on college freshmen, as the first year of college is considered to be a stressful life event due to moving away from home, loss of existing peer group and the challenges of more demanding scholastic endeavors, all potentially resulting in extended psychosocial stress (11, 12). The study was aimed at gathering psychological and biological biomarkers to predict the risk of depression in a sample of students to be followed longitudinally during the freshman year and the following summer. It began prior to the COVID-19 pandemic but was continued (with some modifications) during the first two years of the pandemic, thereby capturing its impact on two freshman cohorts after the onset of the pandemic (2019-2020 and 2020-2021).

It is by now well-established that the COVID-19 pandemic was associated with greater incidence of depression. A large study in the United States looked at depression symptoms in adults before and during the early pandemic period (March and April 2020) and found increased depression symptoms (13). A study in Iceland also found an increase in depression symptoms especially in older adolescent females (14). A meta-analysis in youths found that during the pandemic anxiety and depression symptoms were higher later in the pandemic, and higher in girls (15). There is emerging evidence that in 2020, college freshmen were profoundly affected by COVID-19. During the beginning of the pandemic in China, when sampled at a single time point, the prevalence of anxiety was 23% and the prevalence of depression was 48% (16), with comparable levels observed in US college students (17). The most relevant recent Healthy Minds Report across all college levels showed that 44% of students met criteria for depression (18). However, these studies did not systematically provide comparisons to previous rates of incidence, nor did they carry out any longitudinal follow-up.

Our Michigan Freshmen Study focuses on smaller samples but offers several methodological advantages for parsing the interplay between genetics and environment in vulnerability to depression and anxiety. First, we characterize the subjects at baseline, as they begin their college year, using psychological rating scales, and then use a longitudinal design to follow them throughout the academic year and the following summer. Secondly, we genotype subjects and compute MDD-PRS to analyze its interactions with stress in shaping the emergence of anxiety and depression. Finally, we compare depression and anxiety symptoms in two sequential cohorts during the COVID-19 pandemic to a pre-pandemic cohort. This has enabled us to describe the relative impact of different levels of environmental stress on the development of mood disorders by contrasting the freshman experience alone versus its combination with pandemic stress. By contrast, previously published studies in college students have limited clinical measures and no genetic information (19, 20), or there are no measures pre-COVID-19 (17).

While our primary dependent variables are measures of symptoms of depression and anxiety throughout the freshman year, we also provide our subjects with wearable devices that capture behavioral changes such as physical activity and sleep, as decreased activity and sleep have been implicated in poorer health outcomes, including depression (21, 22). This combination of measures sheds light on the relative role of genetics and environment in shaping the impact of life stressors, including the pandemic, on a young population.

## RESULTS

### Incidence of Anxiety and Depression During the Freshmen Year

A total of 743 subjects were enrolled in this study, and 390 subjects completed the full longitudinal follow-up. Only a subset of these subjects (N=331) could be used for the genetic analyses due to the fact that the MDD-PRS is not appropriate for non-Caucasian subjects (23). This report focuses on this subset of subjects in order to analyze the interplay of genetic and non-genetic variables. A timeline of the study is presented in **Figure 1**. Prior to the pandemic, the study ran from September to September and mood was assessed quarterly (blue arrows). After the pandemic, follow-up was extended to December of the sophomore year, and the frequency of mood testing was increased to monthly assessments (additional red arrows).

**Figure 1.**
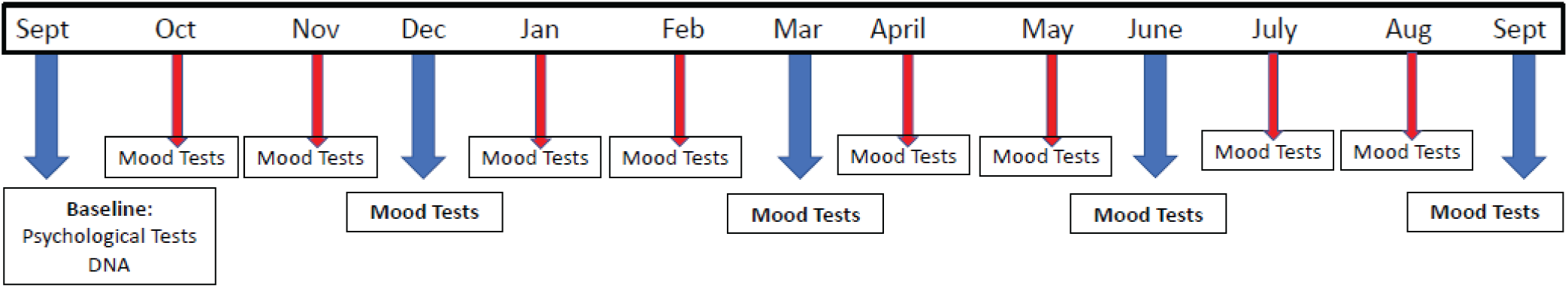
Timeline of Sampling. Baseline had standard questionnaires collected, and these were repeated throughout the year, as indicated by the arrows. Cortisol was also collected at various time points, and DNA was collected at baseline. All questionnaires except the CTQ and NEO PI-R were repeated as Mood Tests throughout the Year. Phase 1 only had blue arrow timepoints. Phase 2 had all blue arrows. In March 2020, several pandemic-related changes happened. March 2020-June 2020, the students were sent home, everyone sheltered-in-place, and remote instruction began. We began monthly mood assessments as indicated by the red arrows. March 2020-October 2020, social distancing was in place at the University of Michigan. A brief shelter-in-place happened again in October 2020. Finally, students were urged to leave campus again in November 2020 and classes went remote until the end of the term. Phase 3 had monthly monitoring with all blue and all red arrows.

We assessed anxiety symptoms by the General Anxiety Disorder-7 (GAD-7) scale, and depression symptoms by the Patient Health Questionnaire-9 (PHQ-9) scale. To determine the incidence of anxiety and depression, we used the highest value at any follow-up and applied the standard cut-off of ≥10 for both GAD-7 and PHQ-9 at baseline and at the 3-, 6-, 9– and 12-month matched time points. **Table 1** shows the percent of subjects who developed depression, anxiety, “Either” depression or anxiety, and “Both” depression and anxiety for each Phase. Phase 1 consisted of the pre 2019 cohort (pre-COVID-19), Phase 2 consisted of the 2019-2020 cohort (the year COVID-19 hit), and Phase 3 consisted of the 2020-2021 cohort (the year after COVID-19). Overall, 26-40% met criteria for either mood disorder during the school year, depending on the stress of the year. Depression incidence at follow-up increased compared to baseline every year. The increases in anxiety symptoms at follow-up were seen only during Phases 1 and 2, as Phase 3 was similar to baseline. However, “Either” anxiety or depression, and “Both” anxiety and depression increased at follow-up every year.

**Table 1.** Incidence of Anxiety, Depression, Either Anxiety or Depression and Both Anxiety and Depression in College Freshmen at the University of Michigan by Cohort.

|  | Phase 1 | Phase 2 | Phase 3 |
| --- | --- | --- | --- |
| **Depression** |  |  |  |
| Baseline | 0.95% | 7.76%<sup>†</sup> | 13.63%<sup>††</sup> |
| Follow-Up | 16.19%<sup>***</sup> | 30.17%<sup>***†</sup> | 33.63%<sup>***††</sup> |
| **Anxiety** |  |  |  |
| Baseline | 4.76% | 8.62% | 15.45%<sup>†</sup> |
| Follow-up | 18.09%<sup>**</sup> | 26.72%<sup>***</sup> | 23.64% |
| **Either** |  |  |  |
| Baseline | 4.76% | 13.79%<sup>†</sup> | 20.91%<sup>†††</sup> |
| Follow-up | 25.71%<sup>***</sup> | 40.52%<sup>***</sup> | 37.27%<sup>*</sup> |
| **Both** |  |  |  |
| Baseline | 0.95% | 2.59% | 8.18%<sup>†</sup> |
| Follow-Up | 8.57%<sup>*</sup> | 16.38%<sup>***</sup> | 20%<sup>*†</sup> |
| **N's** | 105 | 116 | 110 |

†, Compared to Phase 1, $p < 0.05$ , ††, $p < 0.01$ , †††, $p < 0.001$
\*, Compared to Baseline, $p < 0.05$ , \*\*, $p < 0.01$ , \*\*\*, $p < 0.001$

Differences by sex are shown in **Table S1**. For females, all follow-up time points were different from their respective baseline values for Phase 1 for depression and the “Either” condition, with up to 46% of females exhibiting either disorder. Anxiety was only different at follow-up for Phase 2, the year COVID-19 hit. Interestingly, males were only different at follow-up for Phase 1 for either condition.

In comparing Phases 2 and 3 to Phase 1 for all subjects in **Table 1**, differences were observed at baseline for depression and “Either”. For Phase 3 compared to Phase 1, differences were observed only at baseline for anxiety and “Both” conditions. For females, in comparing between Phases 2 and 3 to Phase 1 for depression, differences were observed at baseline (**Table S1**). Additional baseline differences in females were between Phases 1 and 3 for anxiety, “Either” and “Both” conditions. Males did not differ between Phases.

The differences between phases and sex are illustrated in **Figure 2**. **Fig. 2A** shows a clear pattern of increasing baseline PHQ-9 scores at the start of the freshman year, though this increase was largely driven by females (**Fig. 2B-C**). **Fig. S1A-C** shows that GAD-7 scores increased in Phase 3, the first school year that started after the COVID-19 pandemic. However, there were no differences in the highest follow-up GAD-7 scores during the year (**Fig. S1D-F**), suggesting that the pandemic did not further increase anxiety scores during the year. However, follow-up depression scores tell a different story. Depression scores increased in Phase 2 for both males and females and continued to rise in Phase 3 for females but not for males (**Fig. 2D-F**), underscoring sex differences in long-term response to the pandemic.

**Figure 2.**
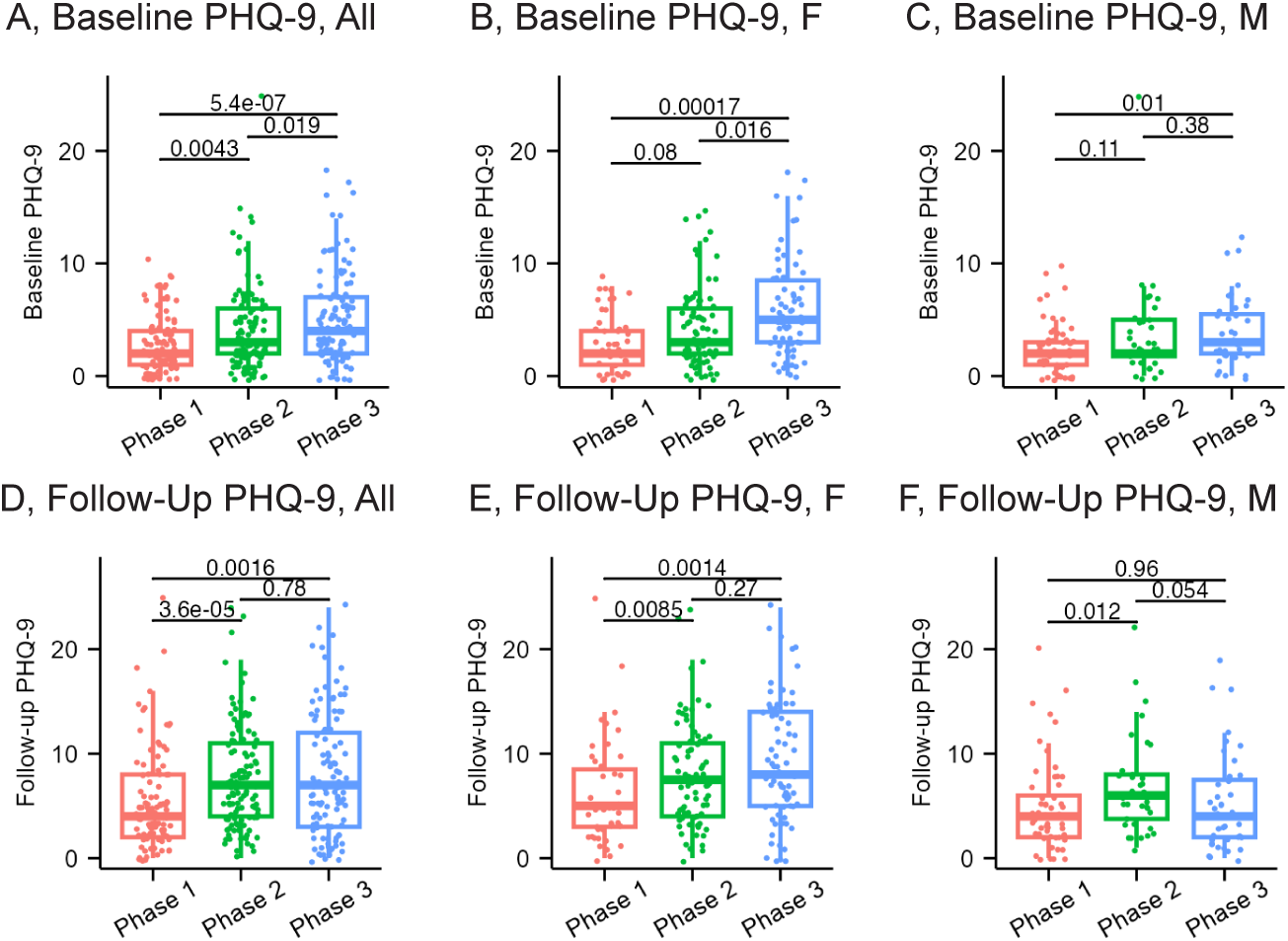
Highest Depression Scores at Baseline and Follow-Up for Each Phase by Sex. <u>For baseline</u>, A) Depression scores were higher in Phase 2 compared to Phase 1, Phase 3 compared to Phase 1, and Phase 3 compared to Phase 2. B-C) The increase was mostly due to females, with males only having higher scores in Phase 2 compared to Phase 1. <u>For follow-up</u>, D) Depression scores were higher in Phase 2 compared to Phase 1, and Phase 3 compared to Phase 1. E-F) The increase was mostly due to females, with males only having higher scores in Phase 2 compared to Phase 1. Medians and IQR. (Phase 1, N=105 (47 Females, 58 Males); Phase 2, N=116 (80 Females, 36 Males); Phase 3, N=110 (71 Females, 39 Males)). Statistical output is shown in **Supplemental Table 4**.

To better illustrate sex differences in depression symptoms during the year, **Fig. S2** shows males compared to females on PHQ-9 scores for Phases 1-3. In Phase 1, there was a significant peak in depression scores in March for females, and females end up higher at the end of the year. In Phase 2, females were increased by May 2020 and remain elevated. In Phase 3, females were significantly higher for the entire year.

### Mood and Activity During the First Year of COVID-19

During Phase 2, which represents the first year of Covid (2019-2020), we collected monthly mood ratings along with collecting daily activity and sleep measures using Fitbits, and we extended these procedures until December 2020 to obtain a more complete picture for this cohort. As shown in **Figure 3A and 3B**, depression and anxiety symptoms significantly increased during COVID-19 (March 2020 and on) relative to ratings in the same cohort prior to the onset of the pandemic. As shown in **Fig. 3C and 3D**, the average number of steps per day decreased dramatically after the onset of COVID-19, and the average minutes asleep per day increased during COVID-19 compared to pre-COVID-19. **Fig. 3E** compares activity in those without depression to those who exhibited depression symptoms at some point during the freshman year. The group that exhibited depression sometime during the course of the year took fewer daily steps prior to the pandemic. But, the striking effect was the dramatic overall decrease in activity in March and April 2020 affect all subjects, followed by limited recovery through the summer and fall months of 2020.

**Figure 3.**
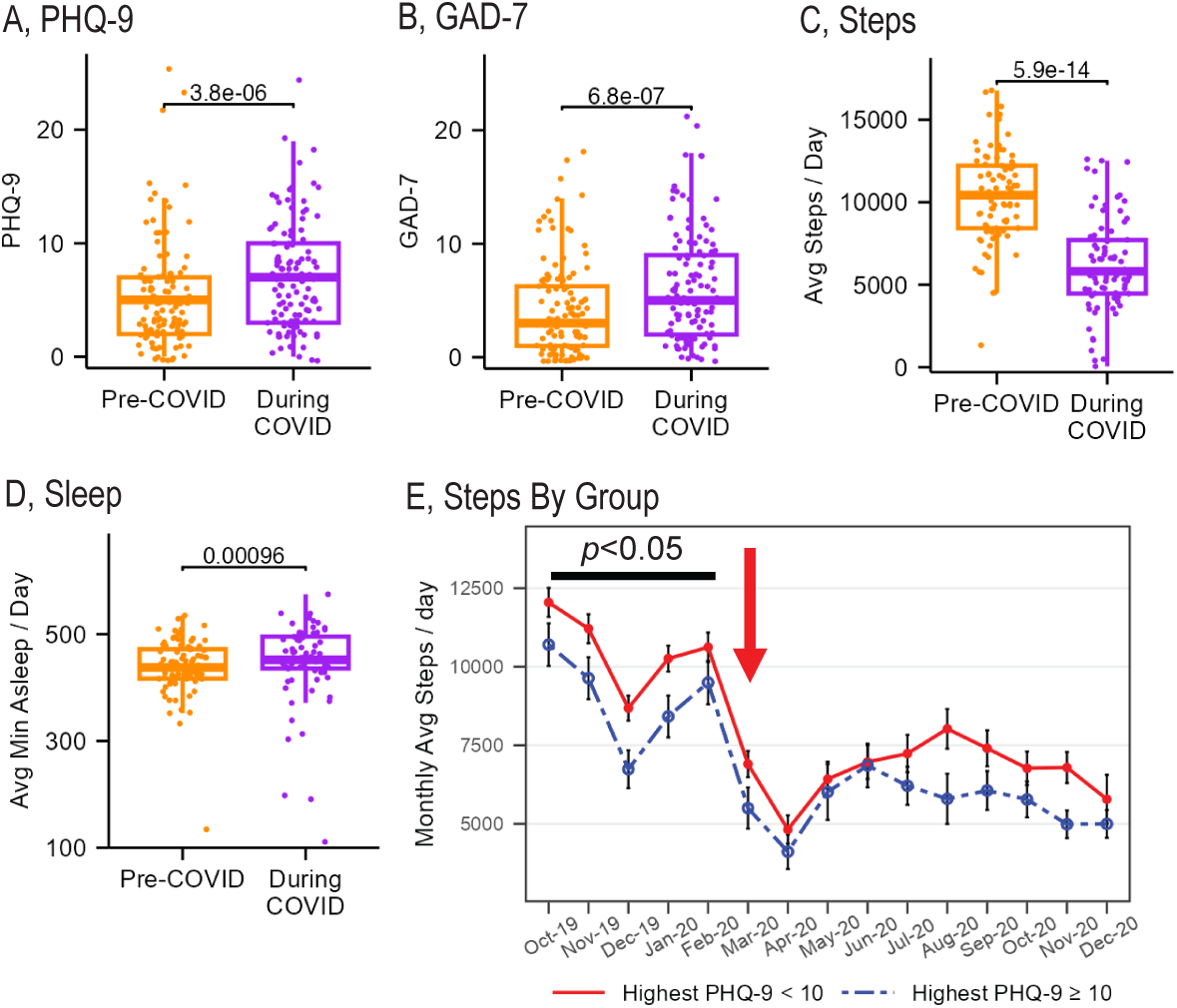
Pre– and During COVID-19 Longitudinal Differences in Mood, Activity and Sleep in the 2019-2020 Cohort, as well as Activity by Month for Depression. B) Depression Scores were higher during COVID-19 compared to pre-COVID-19. B) Anxiety Scores were higher during COVID-19 compared to pre-COVID-19. C) Average number of steps per day were decreased during COVID-19 compared to pre-COVID-19 (*p*=2.7e-11). D) Average sleep per day was increased during COVID-19 compared to pre-COVID-19 (p=7.8e-4). The pandemic led to a dramatic decrease in daily activity across all groups. In addition, activity was modulated by the propensity to develop symptoms of Depression. E) Average number of steps per day were lower at several time points for individuals who develop Depression compared to those without Depression (*F*(1,73.18)=2.996, *p*=0.088), and there was a main effect of time (*F*(14,813.38)=63.695, *p*<0.001), but no interaction (*F*(14,814.13)=1.033, *p*=0.417). If we look at just October through February, the two groups were significantly different *p<0.05. Degrees of freedom using Satterthwaite’s approximation. A-D) Medians and IQR; E) Means and SEMs. (PHQ-9 and GAD-7, N=116; Steps, N=80; Sleep, N=75; Steps No Depression, N=63; Steps, Depression, N=27). Statistical output is shown in **Supplemental Table 5**. Red arrow denotes when COVID-19 hit.

### MDD-PRS and Stress Vulnerability and Resilience

#### Correlation of MDD-PRS with Baseline and Follow-Up Depression

We calculated a polygenic risk score for depression (MDD-PRS) for those subjects that met all criteria for follow-up longitudinal data and ethnicity (N=331). The correlation of this MDD-PRS to depression scores in Phases 1, 2 and 3 is shown in **Figure 4**. MDD-PRS correlated significantly with baseline depression scores in Phases 1 and in Phase 2 (Phase 2 baseline scores were collected prior to COVID-19). MDD-PRS also correlated significantly with follow-up depression scores during Phase 1. However, MDD-PRS was no longer correlated with follow-up depression scores in Phases 2 or 3 –i.e., during the two COVID-19 years. Thus, MDD-PRS correlated with depression under the more typical stress of an academic year (Phase 1), but not under the more complex stress induced by the pandemic (follow-up Phases 2 and 3). **Fig. S3** shows that baseline anxiety was not significantly correlated with the MDD-PRS.

**Figure 4.**
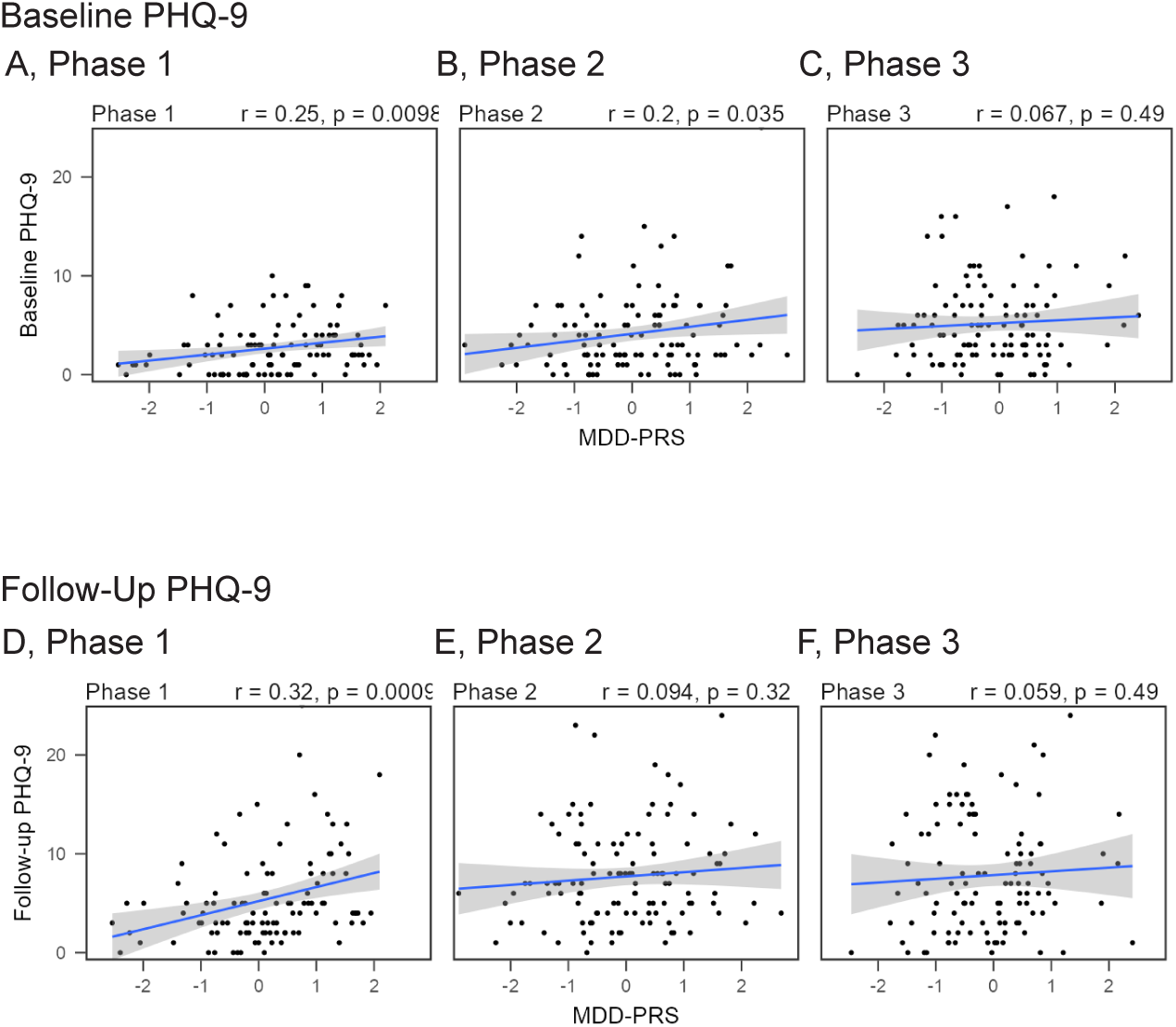
Correlations between MDD-PRS and Baseline and Follow-Up Symptoms of Depression. A-C) For Phases 1 and 2, there were significant correlations at baseline between MDD-PRS and depression symptoms, but not Phase 3. D-F) For Phase 1, there was a significant correlation at follow-up between MDD-PRS and depression symptoms, but not for Phases 2 or 3. (Phase 1, N=105; Phase 2, N=116; Phase 3, N=110).

### Relative Impact of Genetic Risk and Sex on Mood Before and During COVID-19

We then asked how genetic risk modulates stress reactivity across sexes both pre– and during pandemic. We conducted a median split for MDD-PRS and examined the longitudinal pattern of depression and anxiety scores for each sex. We contrasted subjects in the top half of the MDD-PRS distribution (“High MDD-PRS”) to those in the bottom half of the MDD-PRS distribution (“Low MDD-PRS”). **Figure 5** shows the depression scores by month for these two groups for Phases 1, 2 and 3 for both males and females. During Phase 1 (**Fig. 5A and 5D**) the High MDD-PRS group consistently exhibited higher depression scores through the year relative to the Low-MDD-PRS group, especially in females (p <0.01). However, this difference was erased during Phase 2 and Phase 3 (**Fig. 5B-C and 5E-F**). As can be readily seen, the pandemic eliminated the differential impact of genetic predisposition, and the Low MDD-PRS group, across both sexes, showed increased rates of depression and was no longer distinguishable from the High MDD-PRS group (**Fig. 5B and 5E**). In fact, during Phase 3, Low MDD-PRS females showed a trend for higher rates of depression symptoms than High MDD-PRS females (**Fig. 5C**).

**Figure 5.**
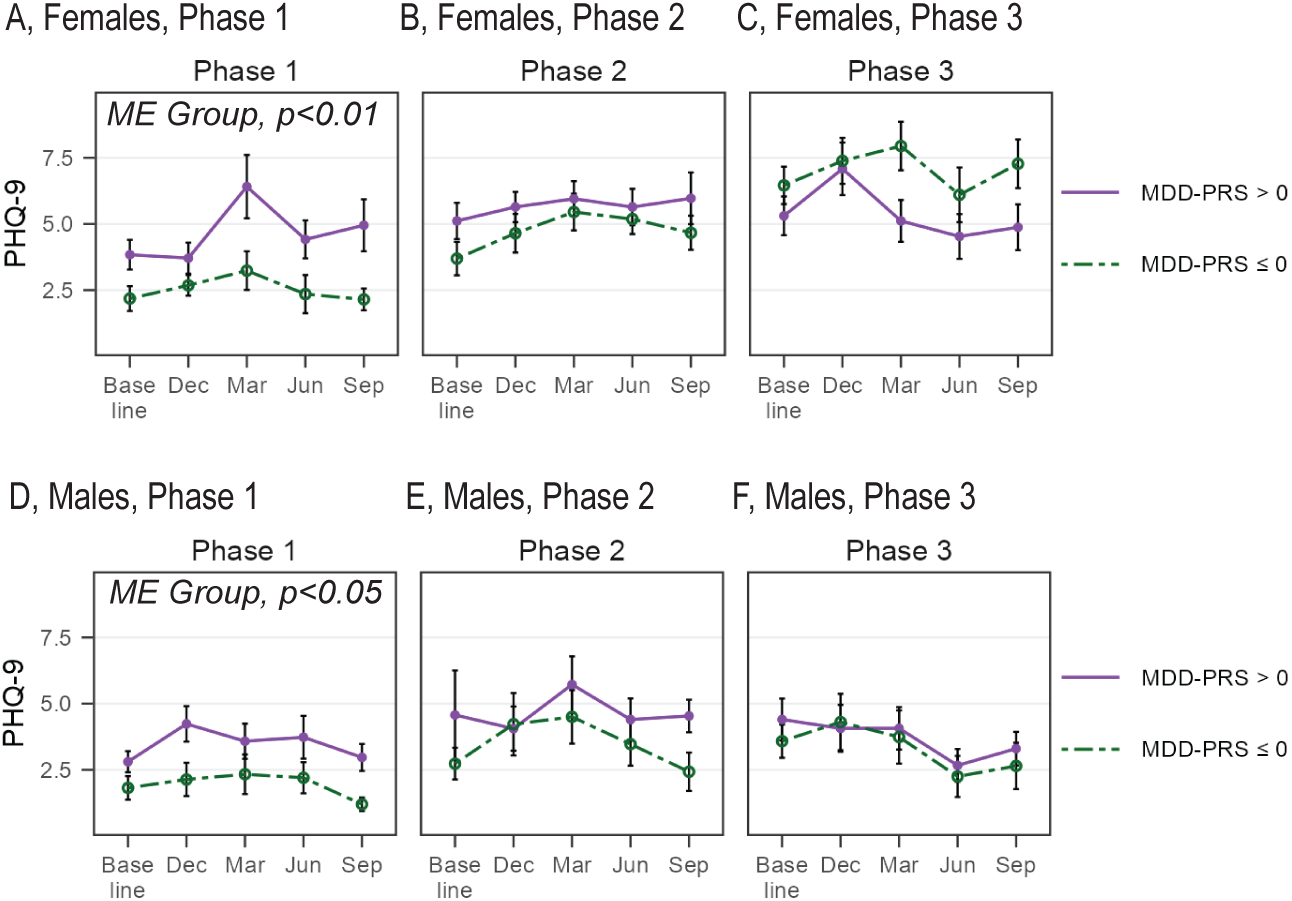
Depression Scores for those with High and Low MDD-PRS across Phases by Sex– Median Split: Monthly Depression Scores (Means and SEMs) in the High MDD-PRS and Low MDD-PRS Groups across study phases. A-C) Depression scores in females over time for High MDD-PRS versus Low MDD-PRS groups, showing higher levels in the High MDD-PRS group in Phase 1, and comparable levels in Phase 2, and a non-significant trend for higher levels in the Low MDD-PRS group in Phase 3. D-F) Depression scores in males over time for High MDD-PRS versus Low MDD-PRS groups, showing higher levels in the High MDD-PRS group in Phase 1, and comparable levels in Phase 2 and 3. Phase 1, Females: Group (*F*(1,45.06)=7.94, *p*<0.01), Time (*F*(4,166.76)=3.33, *p*<0.05), Interaction (*F*(4,166.76)=0.91, *p*=0.46); Phase 1, Males: Group (*F*(1,56.66)=4.81, *p*<0.05), Time (*F*(4,200.96)=2.01, *p*=0.09), Interaction (*F*(4,200.96)=0.47, *p*=0.76); Phase 2, Females: Group (*F*(1,79.11)=1.52, *p*=0.22), Time (*F*(4,277.44)=1.83, *p*=0.12), Interaction (*F*(4,277.44)=0.33, *p*=0.86); Phase 2, Males: Group (*F*(1,39.97)=0.92, *p*=0.34), Time (*F*(4,117.12)=1.28, *p*=0.28), Interaction (*F*(4,117.12)=0.73, *p*=0.57); Phase 3, Females: Group (*F*(1,39.28)=2.69, *p*=0.10), Time (*F*(4,263.95)=3.56, *p*<0.01), Interaction (*F*(4,263.95)=1.75, *p*=0.14); Phase 3, Males: Group (*F*(1,37.28)=0.16, *p*=0.69), Time (*F*(4,128.20)=3.69, *p*<0.01), Interaction (*F*(4,128.20)=0.21, *p*=0.93).

Across sexes, COVID-19 greatly increased the odds of developing depression in the Low MDD-PRS group–– 4.15x higher odds during the first pandemic year (*p*<0.05) and 5.95x higher odds (*p*<0.005) during the second pandemic year. By contrast, the odds of developing depression for the High MDD-PRS group remained constant across all three phases. Thus, while a low genetic risk appeared protective under more typical periods of stress (pre-pandemic), its ability to provide resilience was completely eroded by the pandemic. Parallel analyses on anxiety (**Fig. S4**) show a similar pattern for each sex.

When we collapse across follow-up time points, as shown in **Figure S5**, the Low MDD-PRS females had significantly increased depression scores between Phases 1 and 2 and between Phases 1 and 3. However, the High MDD-PRS females were not different on depression scores between any of the Phases. For males, we did not observe any increase between any of the Phases for either High MDD-PRS or Low MDD-PRS for depression. Thus, in our sample, the increased rate of depression during the pandemic was almost completely attributable to subjects with low genetic risk for MDD, especially females.

### Feature Selection and Principal Component Analysis of Baseline Variables in Predicting Depression

In order to ascertain which baseline psychiatric measures were useful in predicting follow-up PHQ-9, we employed cross-validated recursive feature elimination using a random forest algorithm. This approach was used as several of these measures were highly correlated with each other (24, 25). We included individuals from all Phases together in this analysis and found that out of 16 baseline measures, the following ten measures best explained follow-up depression scores: the Risky Family Questionnaire (RFQ; family history), Childhood Trauma Questionnaire (CTQ; family history), NEO Personality Inventory-Revised (Neuroticism; trait), Spielberger State-Trait Anxiety Inventory (SB Trait; trait) Spielberger State-Trait Anxiety Inventory (SB State; state), Positive and Negative Suicide Ideation Inventory-Positive (PANSI+; state), Positive and Negative Suicide Ideation Inventory-Negative (PANSI-; state), Patient Health Questionnaire-9 (PHQ-9; state), General Anxiety Disorder-7 (GAD-7; state), and Perceived Stress Scale-10 (PSS; state). Due to the high correlation between these measures, we used a principal component analysis to collapse them into fewer dimensions. **Figure 6A** depicts the percent that each of these measures contributes to PCA dimension 1 which explained 58.55% of the total variance. The first PCA dimension strongly correlated with follow-up depression scores, while PCA dimension 2 did not add any explanatory power. In **Figure 6B**, we plotted the subjects along PCA dimension 1 and 2. Subjects who develop depression symptoms have PCA dimension 1 > 0 with an odds ratio of 10.77 (*p*<0.001), showing that individuals higher on PCA dimension 1 were much more likely to develop depression symptoms. For the regression analysis, PCA dimension 1 was transformed into a Z-score and renamed to the baseline Affect Score (AS). **Table 2** shows a linear regression model with heteroskedasticity consistent standard errors with the highest follow-up PHQ-9 score as the dependent variable (26, 27). Both the MDD-PRS and Affect Score (AS; Z-score of Dimension 1) were significant in the overall regression model (Adj. R^2^=46.36%, *F*=41.74, *df*=7,323, *p*<0.001). There was a nonsignificant trend for subjects in Phase 3 to have higher follow-up PHQ-9 scores than those in the earlier Phases. The Sex by Phase 3 interaction also shows that the higher scores in Phase 3 were largely attributable to female subjects, after accounting for the Affect Score and MDD-PRS. The overall model explained 46.36% of the variance in the follow-up depression score. MDD-PRS was significant even after accounting for Affect Score, Phase & Sex. However, Affect Score had a higher explanatory power than MDD-PRS. The regression also showed that the Phase 3 sex difference could not be explained by AS at baseline or MDD-PRS. An alternative model using depression as a categorical variable (shown in **Table S2**), was comparable to the linear model (AUROC = 0.835, **Fig. S6**).

**Figure 6.**
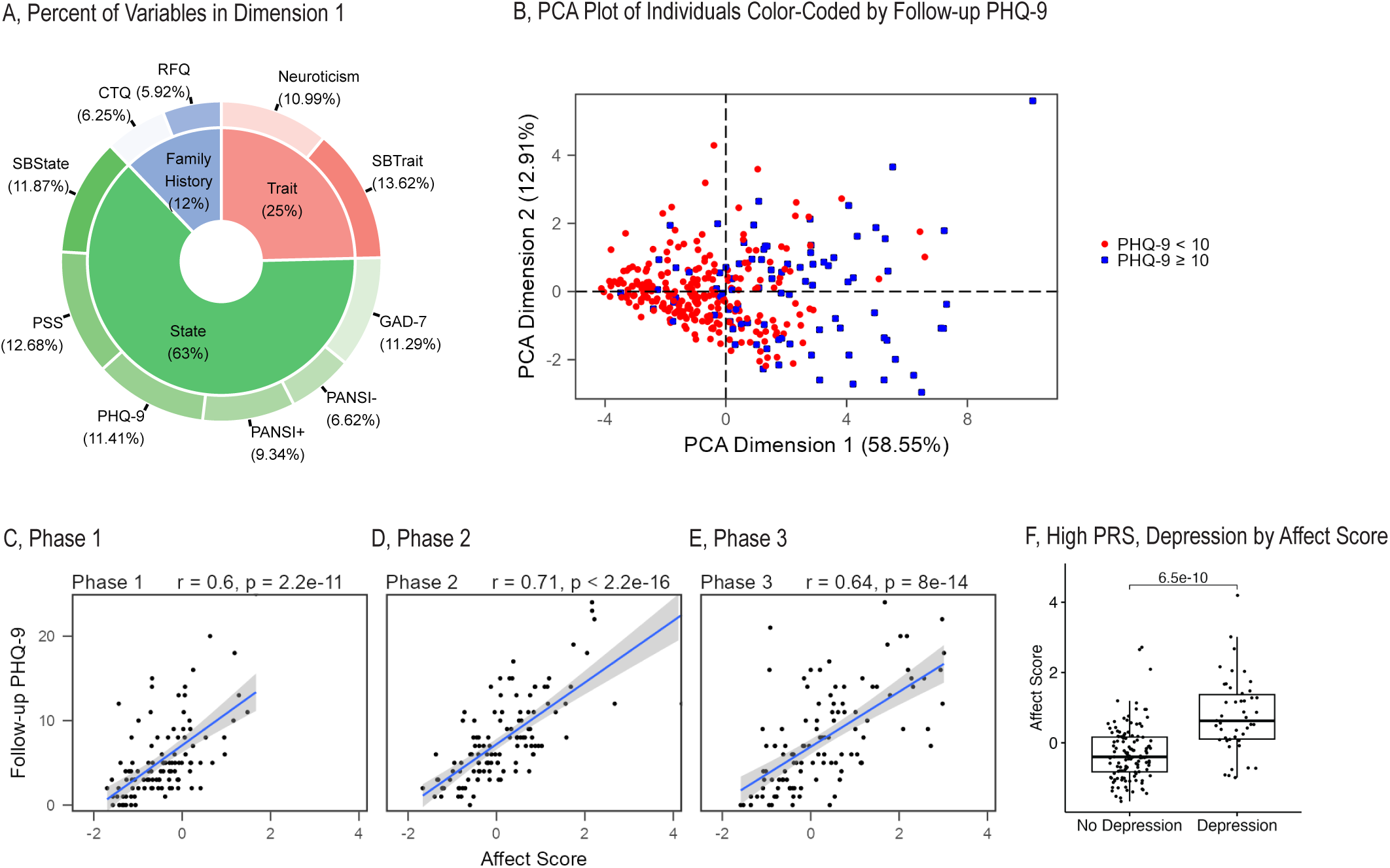
Principal component analysis for variables involved in predicting depression. A) Components of PCA Dimension 1 with percentage contribution to that dimension. PCA Dimension 1 explains 58.55% of the variance of these variables. B) During the year, most subjects that reported a PHQ-9 score of 10 or more had an Affect Score ≥ 0 at baseline (OR = 10.77, p < 0.001). C-E) The Affect Score (scaled PCA dimension 1) highly correlated with follow-up depression scores across all Phases. F) High MDD-PRS with a low Affect Score at baseline were much less likely to become depressed than their counterparts with high Affect Score. (A-B: N=331; C-E: Phase 1, N=105; Phase 2, N=116; Phase 3, N=110; F: No Depression, N=124; Depression, N=46).

**Table 2.** Linear Regression Model with Follow-up Depression Score as Dependent Variable.

|  | PHQ-9 Score |
| --- | --- |
| PRS | 0.521 (0.213) * |
| Sex |  |
| Female | 0.380 (0.743) |
| Affect Score | 3.343 (0.315) *** |
| Phase |  |
| 2 | 0.276 (0.726) |
| 3 | -1.24 (0.716) † |
| Sex x Phase Interaction |  |
| 2 | 0.086 (1.026) |
| 3 | 1.983 (1.081) † |
| Intercept | 6.649 (0.492) ** |
| N's | 331 |

†, $p < 0.1$ \*, $p < 0.05$ \*\*, $p < 0.01$ \*\*\*, $p < 0.001$

As shown in **Figure 6C-E**, the AS was positively correlated with the highest follow-up depression score across all Phases and sexes. However, Affect Score did not correlate with the MDD-PRS (data not shown). This suggests that we have identified a set of baseline psychological variables, including family history, trait and state, that were highly predictive of vulnerability or resilience to future stress, regardless of whether the stress could be classified as typical (freshman year) or highly unusual (pandemic stress).

While our analyses characterized both the existence and the limits of genetic resilience, a remaining question surrounds the basis of resilience in the face of stress, especially in genetically vulnerable individuals. We, therefore, focused on all the subjects with High MDD-PRS who did not become depressed across Phases 1, 2 and 3 and asked whether they have characteristics that distinguish them from those who do. As shown in **Figure 6F**, once again the Affect Score at baseline proved to be a major factor. Thus, individuals with high genetic risk who had a low Affect Score at baseline were much less likely to become depressed than their counterparts with a high Affect Score. It therefore appears that mechanisms that lead to a low Affect Score in the face of high genetic risk are especially important to understand as protective factors against future depression.

## DISCUSSION

This longitudinal study characterized the impact of the COVID-19 pandemic on depression and anxiety states in a cohort of young, healthy subjects facing the stress of entering college and compared them to a previous cohort facing college before the pandemic. Not only did our results highlight the strong impact of the pandemic on this population, but they provided a novel perspective on the interplay between genetic risk and levels of environmental stress in shaping stress reactivity. In particular, our analyses led us to characterize two categories of stress resilience–– “genetic resilience” as indexed by a low biological risk for mood disorders (Low MDD-PRS), and “psychosocial resilience” which is evident in individuals with low Affect Score at baseline. Moreover, Affect Score at baseline proved to be more important than genetic risk in its ability to predict susceptibility or resilience to psychosocial stress.

Our findings support the premise that the freshman year of college represents a significant psychosocial stressor (16-18, 28), with between 26%-40% of freshman developing symptoms that meet criteria for either anxiety or depression using matched time points, depending on the year. The pandemic represented a significantly greater stressor, as we observed the highest levels of negative affect in the 2020-2021 cohort of freshman including measures of depression and anxiety. Interestingly, this effect was mostly due to females. But perhaps the most drastic pandemic-induced change during the first year was the large decrease in physical activity and increase in sleep as monitored through wearable devices. A recent study found that a reduction in self-reported physical activity during COVID-19 was more apparent in individuals who had been more active prior to the pandemic (21), and that decreased activity, in turn, impacted mood.

We then asked how the response to stress was modulated by genetic risk for depression. Prior to the pandemic, depression symptoms at baseline were positively correlated with MDD-PRS. However, under the pandemic, modulation by genetic risk all but disappeared. Indeed, it was the Low MDD-PRS group that exhibited the greater shift during COVID-19, showing more depression symptoms, especially in females. This suggests that the current markers for genetic risk for MDD may not be capturing vulnerability to certain classes of stressors that became especially powerful during the pandemic. A finer analysis of the heterogeneity of MDD, including sex-specific variables (29) might be necessary for enhancing the ability of PRS-MDD to be predictive across populations, sex, and a broader range of stress conditions.

An equally plausible interpretation is that a low genetic propensity to MDD offers a limited measure of “genetic resilience” that is primarily relevant under certain types of (familiar?) stress conditions. Developmental, experiential, and cognitive mechanisms are likely able to supersede genetic factors and provide effective coping strategies against stress, thereby providing “psychological resilience”. In order to get at baseline measures that best predict follow-up PHQ-9, we repeatedly resampled our data hundreds of times and ran a random forest model on those subsets. We then conducted PCA on these measures, and generated a baseline index that was highly correlated with follow-up PHQ-9 scores both prior to and during the pandemic. We have termed the first dimension of the PCA the Affect Score. It consists of personal history, specific trait anxiety and neuroticism measures, along with state measures at baseline which represent the largest predictive elements of this dimension. Thus, the affective state of the individual at baseline, even when the ratings are below the clinical threshold, appears especially powerful at predicting future propensity for developing significant mood symptoms through the course of the year.

The predictive power of the Affect Score in our sample was notable in the context of exploring psychological resilience in the face of genetic vulnerability. To this end, we compared subjects with High MDD-PRS who developed depression (as might be predicted based on their genetics) to subjects with High MDD-PRS who never developed depression. The two groups were matched in terms of genetic risk, but were highly different on their baseline Affect Score, suggesting the existence of powerful countervailing variables that are protective in the face of biological vulnerability. It will be important to replicate these findings in an independent sample of subjects. This would then represent a starting place for uncovering biological, cognitive and/or social mechanisms that impart psychological resilience, especially in higher risk populations. An example would be epigenetic changes induced by maternal characteristics that could contribute to the presence of psychological susceptibility or resilience under more severe stress (30).

A notable finding is the disproportionate impact of the pandemic on females in our study, whereby the difference in incidence between females and males was amplified from the years prior to the pandemic to the second year of the pandemic, as females increased 100% and males only increased 50%. Especially surprising was our observation that females with low genetic risk showed the greatest increase in the incidence of depression during the pandemic, and the baseline PHQ9 scores for females with low genetic risk increased significantly and progressively across the phases of the study. The reasons are unclear and are worthy of further investigation. One hypothesis is that while females with high genetic risk may have developed counterregulatory coping mechanisms that served to protect them, those with low genetic risk may have lacked these coping tools. If true, this points to the importance of preventive treatments to induce psychological resilience, especially in this population whose genetic resilience fails to protect them under more extreme stress.

In summary, our work highlights the profound impact of the pandemic on a young healthy population, with the emergence of symptoms of clinical anxiety and depression coupled with a dramatic decrease in physical activity. It sheds light on the interplay between genetic risk and environmental stressors in shaping affective responses, by identifying the existence of “genetic resilience” that appears protective under moderate stress but failed to be protective under the pandemic, and “psychological resilience” that can counteract genetic susceptibility. This lays the groundwork for defining parameters for susceptibility and resilience across different populations and devising targeted interventions to enhance stress resilience.

## METHODS

### Subjects

Subjects were college freshmen at the University of Michigan recruited via email, Facebook and posted flyers by the beginning of their freshman year. Because of COVID-19, the study has been split into three parts: Phase 1 consists of the 2015-2019 cohort, Phase 2 (the year COVID-19 hit) consists of the 2019-2020 cohort, and Phase 3 consists of the 2020-2021 cohort (the year after COVID-19 hit).

The study design during the pre-COVID era included students from multiple ethnic backgrounds, and assessment of several biological variables, e.g., stress blood measures and a laboratory stress test, most of which had to be discontinued during the pandemic. A total of 677 subjects were initially recruited for genotyping. Of these, a total of 509 subjects were recruited for the follow-up portion of the study across all three Phases (i.e., 197, 147 and 165, respectively). Since the current analysis focuses on the impact of the pandemic and its interaction with genetic risk on mood outcomes, and since MDD-PRS was derived from individuals with European ancestry, it was necessary to confine our analysis to the Caucasian subset of subjects. Thus, Phase 1 included N=105 Caucasians, Phase 2 included N=116 Caucasians, and Phase 3 included N=110 Caucasians. COVID-19 represents an important variable for Phase 2, as starting in March 2020, all subjects were sent home from college. All subjects provided written informed consent after receiving a complete oral description of the study.

### Data and Sample Collection

#### Phase 1

Subjects aged 18-19 were enrolled by recruitment after matriculation. Starting in August or September (baseline) they were given the following written 14 questionnaires (16 measures): NEO Personality Inventory-Revised (NEO PI-R), Risky Family Questionnaire (RFQ), Childhood Trauma Questionnaire (CTQ), General Anxiety Disorder-7 (GAD-7), Spielberger State-Trait Anxiety Inventory (SB State, SB Trait), Positive and Negative Suicide Ideation Inventory (PANSI+, PANSI-), Patient Health Questionnaire-9 (PHQ-9), and the Morningness-Eveningness Questionnaire (MEQ), as well as the Perceived Stress Scale-10 (PSS), Multidimensional Scale of Perceived Social Support (MSPSS), Connor-Davidson Resilience scale (CD-RISC), 5-item Dispositional Positive Emotions subscale (Compassion), Pearlin Mastery scale (Mastery), and the 3-item Revised UCLA Loneliness scale (Loneliness).

At 3-, 6-, 9– and 12-months, the PHQ-9, GAD-7, and PANSI were repeated in Qualtrics. At 4-, 8-, and 12-months, the PSS, MSPSS, CD-RISC, Compassion, Mastery, and Loneliness were repeated by paper and pencil. A highly skilled psychiatric nurse conducted both the Structured Clinical Interview for DSM-Non-patients (SCID-NP) and the Family History Method for Research Diagnostic Criteria (FH-RDC) on each subject at the start of the study, and a LIFE interview at the one-year follow-up. All subjects wore Fitbits (Charge HR, 2 or 3) for sleep and activity tracking. For sleep and activity, subjects were included if there was data on at least 25% of the days.

#### Phases 2 and 3

The design was similar to Phase 1 with the following exceptions: Questionnaires were moved to a RedCap database for the September baseline. Starting in May 2020, subjects were asked to complete the repeating questionnaires monthly, including adding a COVID-10 questionnaire; and data collection time was extended until December for each year. The questionnaires were every month starting in September for Phase 3, the 2020-2021 group. When we calculated incidence or depression scores by Phase or Sex, we used the four matched time points across all the Phases.

#### For all Phases

the main follow-up measures were GAD-7 for anxiety, and PHQ-9 for depression at matching time points (at 3-, 6-, 9– and 12-months). The highest follow-up value was used to determine the presence of anxiety or depression during the school year. For classification purposes, we used cut-off values of 10 as these were shown to have high sensitivity and specificity for both PHQ-9 and GAD-7 (31, 32). The number of subjects is lower than those from initial genotyping, due to quality control and filtering of the genetic data, as well as the fact that any subject missing more than two (50%) of the follow-up time points were excluded from the analysis.

### Sample Collection and Processing

Blood samples were collected for genomic DNA at baseline in EDTA BD vacutainer tubes and were processed in the lab by trained investigators using established protocols (Gentra Puregene Blood kit). During the pandemic, we shifted to collecting salivary samples using DNA Genotek tubes (OGR-500). The saliva was processed by the Central Biorepository at the University of Michigan. During Phase 1, we also collected blood, saliva and hair samples at baseline and hair or saliva samples at follow-up time points for endocrine measures. However, endocrine sample collection was disrupted by the pandemic, and these measures will not be discussed in this paper.

### Genotyping and Quality Control

The genotyping was performed at the University of Michigan’s Advanced Genomics Core. For Phase 1, DNA was genotyped on Illumina HumanExome OmniExpress-24 v1.2 or v1.3 chips (N=192). For Phases 2 and 3, DNA was genotyped on Illumina Human Infinium CoreExome-24+ v1.3 chips (N=420). Since there were two different chips used in Phase 1, we needed to update the strand and position script. We took the original bed stem from the v1.3 chip and then downloaded the v1.2 strand file from Dr. Will Rayner’s website at Oxford https://www.well.ox.ac.uk/~wrayner/strand/ and followed his workflow to “updatestrandofpedfile.txt” to generate the new file. Since this also flipped the strand to all forward, we flipped the v1.2 file as well. We identified the common SNPs in R v4.2.3 and merged the two files in PLINK (v1.9). We followed previously published methods and workflows for quality checking and LD-based pruning in PLINK: The sample call rate was 95%, the SNP call rate was 99%, the Minor Allele Frequency was 0.005, the Hardy-Weinberg equilibrium was 0.00001 and the LD-based pruning window size was 50 with a step size of 5 (10). In order to get a combined PRS for all the subjects, we first imputed the genotyping data using the 1000 Genomes Phase 3 reference panel (33). The data was imputed using Minimac 4 on the Michigan Imputation Server (https://imputationserver.sph.umich.edu/index.html). (10). Our population stratification is shown in an MDS plot in **Figure S7**. This plot was generated by combining our data with that of the 1000 genomes project, and we included all of our subjects that self-identified as of European Ancestry (own-Eur).

### MDD-PRS Calculation

Using an additive model, we merged the imputed data and PRSice v.2.3.3 was used for the MDD-PRS calculation using the clumping and thresholding method, with the exception that a secondary analysis was not performed (10). We did not threshold on *p*-value and instead used all SNPs to calculate the PRS. We relied on the most recent MDD GWAS summary statistics including 23andMe (5). After imputation and merging, we had 47,099,827 SNPs. After filtering on MAF (0.1) and quality score of 0.9 and including SNPs which were a part of the GWAS, we were left with 2,705,770 SNPs. After clumping, the PRS was calculated using 61,586 SNPs (34–37). The MDD-PRS was then mean-centered and scaled to a standard deviation (s.d.) of 1. The filtering process and quality control results by Phase are shown in **Table S3**.

### Statistical analyses

All statistical analyses were performed in R v4.2.3. A median split was used to classify subjects as High or Low MDD-PRS. For the analysis of incidence, a Fisher’s Exact test was used. For the analysis of Phase effects on anxiety or depression symptoms alone or separated by sex or PRS, a Mann-Whitney test was performed for unpaired tests, and Wilcoxon signed rank test was used for paired tests. It was also used to test for differences between groups (Pre-COVID vs. During COVID, No Depression vs. Depression) on measures of sleep, activity, or mood. To check for differences over time, a random effects ANOVA model was used with pre-specified contrasts at each time point (38–40). Pearson correlations were run to assess associations between PRS, mood or other data. Odds ratios that were calculated with corrected for small sample sizes and Yate’s continuing correction was applied to the chi-square *p*-values. Either PHQ-9 highest follow-up score or GAD-7 highest follow-up score was used as the dependent variable.

To assess the explanatory power of baseline psychiatric measures on follow we asked which of these measures best explained follow-up PHQ-9. Using repeated cross-validation (repeatedly resampling the data into train / test sets), we applied a random forest algorithm to find the baseline surveys which best predicted the follow-up PHQ-9 score (24, 25). The random forest method was chosen as there was a high correlation among the baseline surveys thereby making a stepwise linear model inappropriate. Once we had the baseline surveys best predicting follow-up PHQ-9, we conducted Principal Component Analysis (PCA) on them to construct the Affect Score which is the Z-score of the first dimension of the PCA (41). The Affect Score was included in the regression analysis with the follow-up PHQ-9 score as the dependent variable and Sex, PRS and Affect Score as the independent variables. A dummy variable for Phase was also included in the model to disentangle the effect of the COVID-19 pandemic, as well as the interaction of sex with this dummy variable. A second model using depression as a categorical variable was also performed. The code will be deposited in github (url link), the SNP data will be deposited in dbSNP (SUB#), and the associated meta-data will be deposited in the University of Michigan Library’s Deep Blue Data Repository after acceptance.

## Data Availability

Data produced in the present study will be made available following publication in a peer-reviewed journal upon reasonable request to the authors.

## Acknowledgments

This study was supported by the Office of Naval Research (ONR) Grants N00014-09-1-0598, N00014– 12-1-0366 and N00014-19-1-2149, the Hope for Depression Research Foundation, and the Pritzker Neuropsychiatric Disorders Research Consortium Fund LLC (http://www.pritzkerneuropsych.org). We would like to thank the Michigan Institute for Clinical and Health Research for help with this study (UL1TR002240).

## Financial Considerations

This work was partially supported by the Pritzker Neuropsychiatric Disorders Research Fund L.L.C. The authors are members of the Pritzker Neuropsychiatric Disorders Research Consortium, which is supported by the Pritzker Neuropsychiatric Disorders Research Fund L.L.C. A shared intellectual property agreement exists between this philanthropic fund and the University of Michigan, Stanford University, the Weill Medical College of Cornell University, the University of California at Irvine, and the HudsonAlpha Institute for Biotechnology to encourage the development of appropriate findings for research and clinical applications. The authors declare no biomedical financial interests and no conflicts of interest.

