## Supplemental Material for "Stress, Genetics and Mood: Impact of COVID-19 on a College Freshman Sample"

**Supplemental Table 1.** **Incidence of Anxiety, Depression, Either Anxiety or Depression and Both Anxiety and Depression in College Freshmen at the University of Michigan by Cohort and Sex.**

Female	Male

Phase 1	Phase 2	Phase 3	Phase 1	Phase 2	Phase 3

**Depression**

Baseline	0%	10%†	17%††	2%	3%	8%

Follow-Up	21%**	35%***	42%**†	12%	19%	18%

**Anxiety**

Baseline	9%	7%	21%‡	2%	11%	5%

Follow-Up	26%	30%***	30%	12%	19%	13%

**Either**

Baseline	9%	15%	28%†	2%	11%	8%

Follow-Up	36%**	46%***	46%*	17%**	28%	21%

**Both**

Baseline	0%	2%	10%†	2%	3%	5%

Follow-Up	11%	19%**	25%*	7%	11%	10%

**N’s**	47	80	71	58	36	39

†, Compared to Phase 1, p<0.05, ††, *p*<0.01

‡, Phase 2 compared to Phase 3, *p*<0.05

*, Compared to Baseline, p<0.05, **, *p*<0.01, ***, *p*<0.001

**Supplemental Table 2. Logistic Regression Model with Incidence of Depression as Dependent Variable.**

Depression

PRS	0.260 (0.152) †

Sex

Female	0.456 (0.589)

Affect Score	1.390 (0.193) ***

Phase

2	-0.014 (0.664)

3	-0.219 (0.659)

Sex x Phase Interaction

2	0.144 (0.815)

3	0.468 (0.817)

Intercept	-1.660 (0.439) **

N’s	331

†, *p*<0.1

*, p<0.05

**, *p*<0.01

***, p<0.001

**Supplemental Table 3. Quality Control and Filtering of Genotyping Data by Phase.**

Phase 1	Phase 2	Phase 3

Initial SNPs	706,628	595,427	595,427

Initial Subjects	257	216	204

Genotype Filter	705,580	594,192	577,545

Call Rate Subjects	255	206	202

Sex Check Subjects	248	204	202

Autosomal SNPs	686,280	575,673	560,469

MAF Filtering	645,085	313,261	305,396

HWE Filtering	644,944	313,113	305,300

*Note: In Phase 1, additional subjects not part of this study were included in the genotyping, filtering QC and imputation process. They were removed after these steps from this study.*

**Supplemental Table 4. Descriptive Statistics for Figure 2 and Supplemental Figure 1.**

| **Timepoint** | **Measure** | **Sex** | **Phase** | **Mean** | **Median** | **StdDev** |
| --- | --- | --- | --- | --- | --- | --- |
| Follow-up | PHQ-9 | Male | Phase1 | 4.86 | 4 | 4.24 |
| Follow-up | PHQ-9 | Female | Phase1 | 6.09 | 5 | 4.96 |
| Follow-up | PHQ-9 | Both | Phase1 | 5.41 | 4 | 4.59 |
| Follow-up | PHQ-9 | Male | Phase2 | 6.78 | 6 | 4.64 |
| Follow-up | PHQ-9 | Female | Phase2 | 8.13 | 7.5 | 4.95 |
| Follow-up | PHQ-9 | Both | Phase2 | 7.71 | 7 | 4.88 |
| Follow-up | PHQ-9 | Male | Phase3 | 5.23 | 4 | 4.78 |
| Follow-up | PHQ-9 | Female | Phase3 | 9.21 | 8 | 5.85 |
| Follow-up | PHQ-9 | Both | Phase3 | 7.80 | 7 | 5.80 |
| Follow-up | GAD-7 | Male | Phase1 | 4.74 | 4 | 4.25 |
| Follow-up | GAD-7 | Female | Phase1 | 6.60 | 5 | 4.67 |
| Follow-up | GAD-7 | Both | Phase1 | 5.57 | 4 | 4.52 |
| Follow-up | GAD-7 | Male | Phase2 | 5.61 | 4 | 4.34 |
| Follow-up | GAD-7 | Female | Phase2 | 7.15 | 6 | 4.97 |
| Follow-up | GAD-7 | Both | Phase2 | 6.67 | 5.5 | 4.82 |
| Follow-up | GAD-7 | Male | Phase3 | 4.18 | 3 | 4.49 |
| Follow-up | GAD-7 | Female | Phase3 | 7.00 | 6 | 5.37 |
| Follow-up | GAD-7 | Both | Phase3 | 6.00 | 5 | 5.24 |
| Baseline | PHQ-9 | Male | Phase1 | 2.43 | 2 | 2.31 |
| Baseline | PHQ-9 | Female | Phase1 | 3.06 | 2 | 2.65 |
| Baseline | PHQ-9 | Both | Phase1 | 2.71 | 2 | 2.48 |
| Baseline | PHQ-9 | Male | Phase2 | 3.69 | 2 | 4.33 |
| Baseline | PHQ-9 | Female | Phase2 | 4.33 | 3 | 3.64 |
| Baseline | PHQ-9 | Both | Phase2 | 4.13 | 3 | 3.86 |
| Baseline | PHQ-9 | Male | Phase3 | 3.90 | 3 | 3.06 |
| Baseline | PHQ-9 | Female | Phase3 | 5.87 | 5 | 4.30 |
| Baseline | PHQ-9 | Both | Phase3 | 5.17 | 4 | 4.00 |
| Baseline | GAD-7 | Male | Phase1 | 2.34 | 2 | 2.45 |
| Baseline | GAD-7 | Female | Phase1 | 3.53 | 3 | 3.31 |
| Baseline | GAD-7 | Both | Phase1 | 2.88 | 2 | 2.91 |
| Baseline | GAD-7 | Male | Phase2 | 2.83 | 1 | 3.94 |
| Baseline | GAD-7 | Female | Phase2 | 3.54 | 2 | 3.81 |
| Baseline | GAD-7 | Both | Phase2 | 3.32 | 2 | 3.85 |
| Baseline | GAD-7 | Male | Phase3 | 4.10 | 3 | 3.63 |
| Baseline | GAD-7 | Female | Phase3 | 5.45 | 4 | 4.91 |
| Baseline | GAD-7 | Both | Phase3 | 4.97 | 4 | 4.53 |

**Supplemental Table 5. Descriptive Statistics for Figure 3.**

| **COVID time** | **Measure** | **Mean** | **Median** | **StdDev** |
| --- | --- | --- | --- | --- |
| Pre-COVID | PHQ-9 | 5.38 | 5.00 | 4.62 |
| Pre-COVID | GAD-7 | 4.30 | 3.00 | 4.13 |
| Pre-COVID | Avg. Steps | 10302.08 | 10436.15 | 2816.22 |
| Pre-COVID | Avg. Min. Asleep | 434.05 | 437.69 | 67.08 |
| During COVID | PHQ-9 | 7.06 | 7.00 | 4.62 |
| During COVID | GAD-7 | 6.22 | 5.00 | 4.84 |
| During COVID | Avg. Steps | 6169.61 | 5814.18 | 2856.32 |
| During COVID | Avg. Min. Asleep | 446.29 | 451.98 | 76.88 |

**Supplemental Figure 1. Highest Anxiety Scores at Baseline and Follow-Up for Each Phase by Sex.** For baseline, A) Anxiety scores were significantly higher in Phase 3 compared to Phase 1 and in Phase 3 compared to Phase 2. B-C) There were differences between Phase 3 and Phase 2 for females, and between Phase 3 and Phase 1, and between Phase 3 and Phase 2 for males. For follow-up, D) Anxiety scores were not significantly different between Phases. E-F) There were no sex differences between Phases. Medians and IQR. (Phase 1, N=105; Phase 2, N=116; Phase 3, N=110).

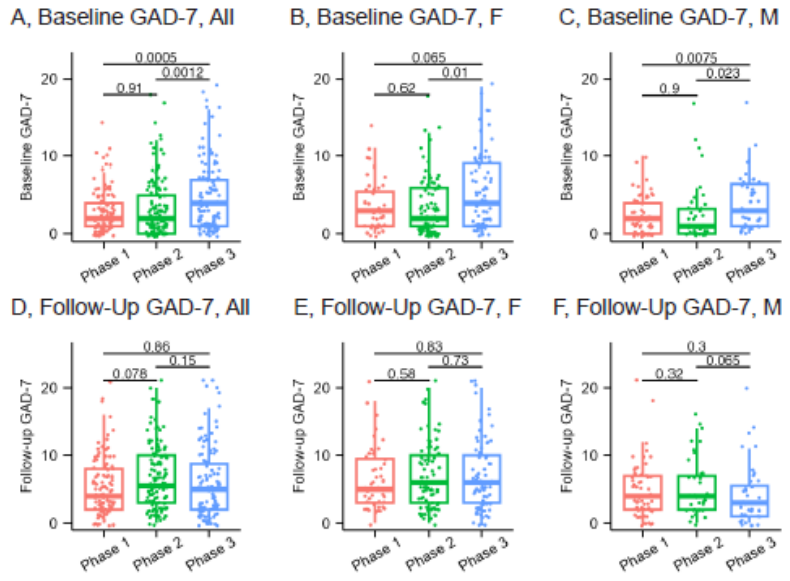

**Supplemental Figure 2. Male and Female Depression Scores Across the Year for Each Phase.** A) In Phase 1, females peaked in March (Sex: *F*(1,105)=2.39, *p*=0.12; Time: *F*(4,376)=4.08, *p*=0.003; Interaction: *F*(4,376)=2.04, *p*=0.088). B) In Phase 2, females were higher starting in May 2020 and remain higher throughout the year. (Sex: *F*(1,112)=3.49, *p*=0.06; Time: *F*(10,978)=2.10, *p*=0.02; Interaction: *F*(10,978)=0.31, *p*=0.98). C) In Phase 3, females were higher throughout the entire year. (Sex: *F*(1,108)=11.07, *p*=0.001; Time: *F*(15,1475)=5.08, *p*<0.001; Interaction: *F*(15,1475)=0.6, *p*=0.87). Means ­± S.E.M. **p*<0.05, ***p*<0.01, adjusted using FDR correction. (Phase 1: Females N=47, Males N=58; Phase 2: Females N=80, Males N=36; Phase 3: Females N=71, Males N=39). Red arrow denotes when COVID-19 hit.

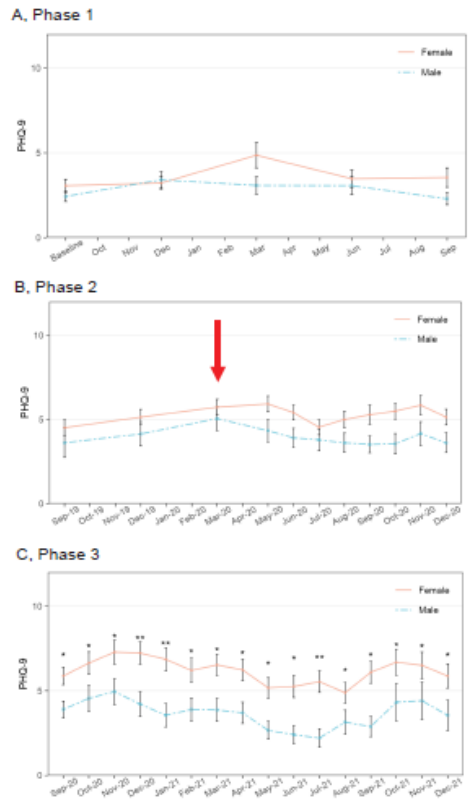

**Supplemental Figure 3.** **Correlations between MDD-PRS and Follow-Up Symptoms of Anxiety.** A-C) There were no significant correlations between PRS and baseline anxiety at any Phase. D-F) There was a significant correlation between PRS and follow-up anxiety for Phase 1, but not for any other Phase. (Phase 1, N=105; Phase 2, N=116; Phase 3, N=110).

**A-C**

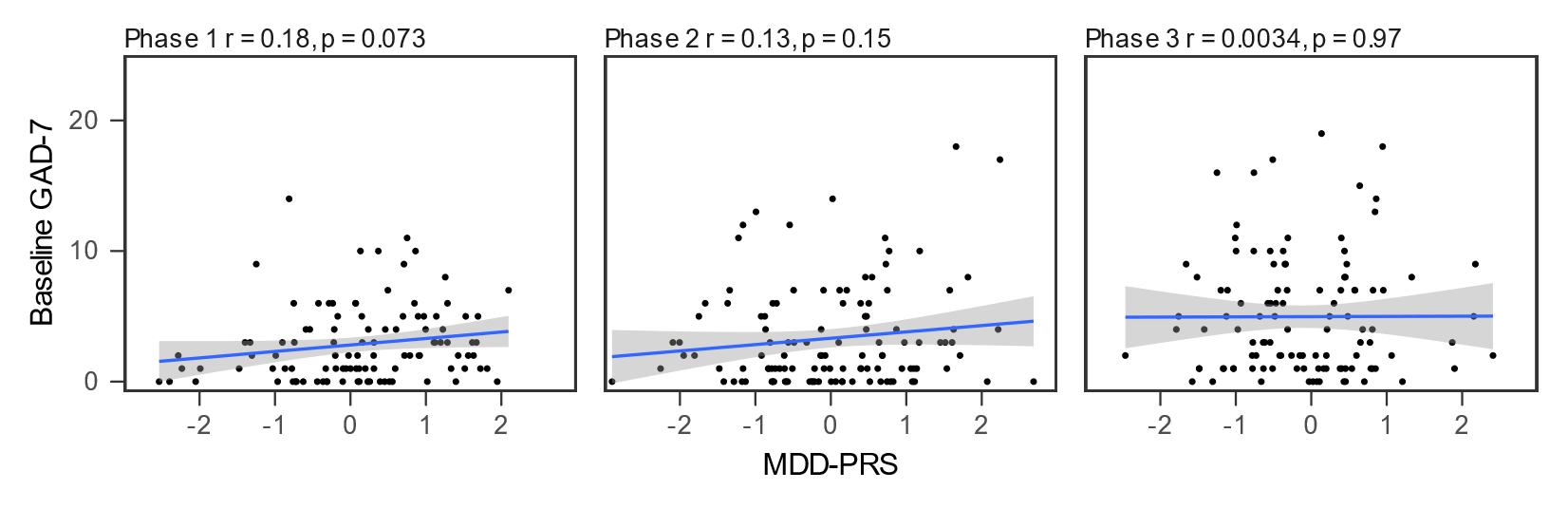

**D-F**

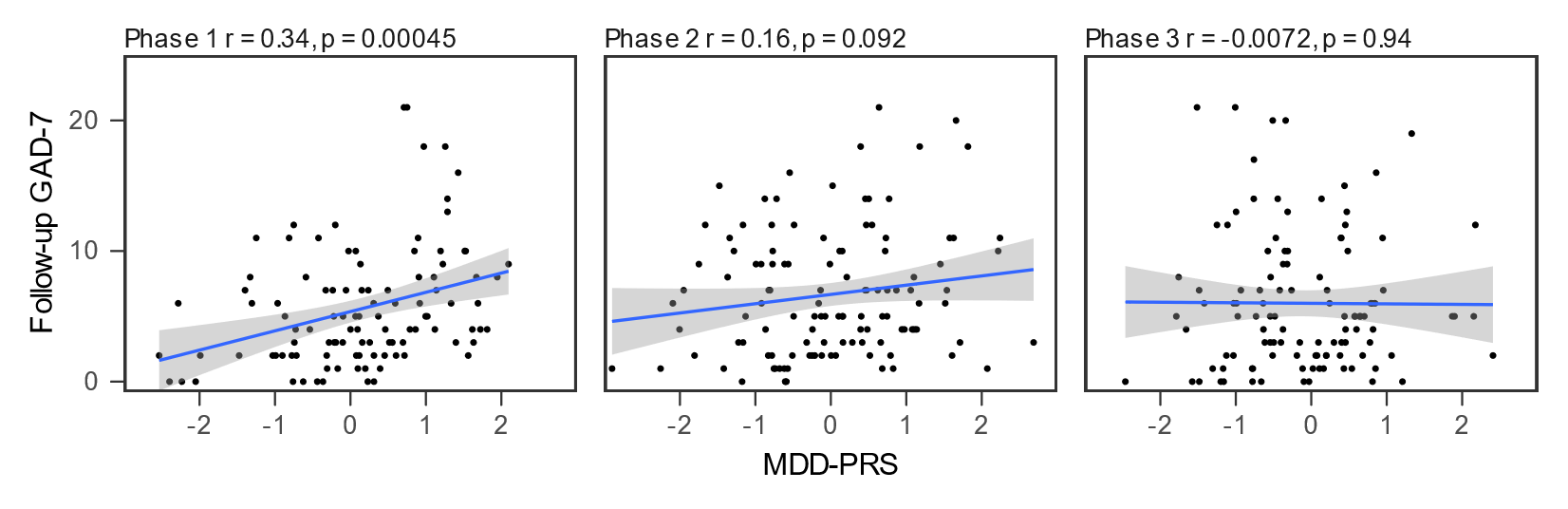

**Supplemental Figure 4. Anxiety Scores for those with High and Low MDD-PRS across Phases by Sex- Median Split:** Monthly Anxiety Scores in the High MDD-PRS and Low MDD-PRS Groups across study phases. A-C) Anxiety scores in females over time for High and Low MDD-PRS groups, showing higher levels in the High MDD-PRS group in Phases 1 and 2, but higher levels in the Low MDD-PRS group in Phase 3. D-F) Anxiety scores in males over time for High and Low MDD-PRS groups, showing higher levels in the High MDD-PRS group in Phase 1, and comparable levels in Phase 2 and 3. Means and SEMs. Phase 1, Females: Group (*F*(1,45.52)=8.32, *p*<0.01), Time (*F*(4,163.78)=3.80, *p*<0.01), Interaction (*F*(4,163.78)=0.62, *p*=0.64); Phase 1, Males: Group (*F*(1,56.47)=3.22, *p*=0.08), Time (*F*(4,198.82)=1.32, *p*=0.26), Interaction (*F*(4,198.82)=0.47, *p*=76); Phase 2, Females: Group (*F*(1,79.04)=4.25, *p*<0.05), Time (*F*(4,275.17)=4.23, *p*<0.005), Interaction (*F*(4,275.17)=0.67, *p*=0.61); Phase 2, Males: Group (*F*(1,32.73)=0.07, *p*=0.80), Time (*F*(4,117.71)=0.98, *p*=0.42), Interaction (*F*(4,117.71)=0.41, *p*=0.80); Phase 3, Females: Group (*F*(1,69.31)=2.01, *p*=0.16), Time (*F*(4,263.79)=0.66, *p*=0.62), Interaction (*F*(4,263.79)=0.74, *p*=0.56); Phase 3, Males: Group (*F*(1,37.42)=0.05, *p*=0.82), Time (*F*(4,128.13)=6.70, *p*<0.001), Interaction (*F*(4,128.13)=0.12, *p*=0.97).

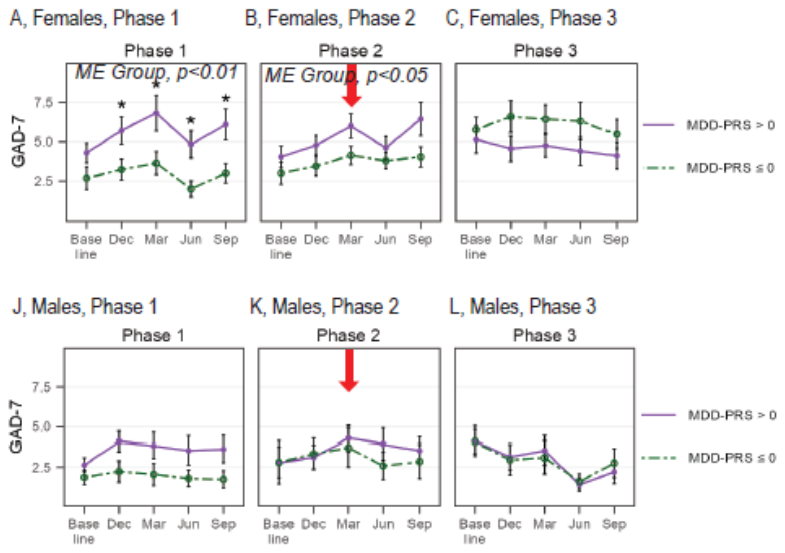

**Supplemental Figure 5. Role of Developing Depression and Anxiety for those with High and Low MDD-PRS across Phases by Sex - Median Split.** A-B) The highest follow-up depression score in females was only significantly different between Phases in the Low MDD-PRS, especially between Phases 1 and 2, and between Phases 1 and 3. C-D) The highest follow-up anxiety score in females was only significantly different in the Low MDD-PRS group between Phases 1 and 3. E-F) The highest follow-up depression score in males was not significantly different between Phases for either MDD-PRS group. G-H) The highest follow-up anxiety score in males was not significantly different between Phases for either MDD-PRS group. Medians and IQR. (Phase 1, High MDD-PRS Female N=25; Phase 1, High MDD-PRS Male N=36; Phase 1, Low MDD-PRS Female N=22; Phase 1, Low MDD-PRS Male N=22; Phase 2, High MDD-PRS Female N=40; Phase 2, High MDD-PRS Male N=18; Phase 2, Low MDD-PRS Female N=40; Phase 2, Low MDD-PRS Male N=18; Phase 3, High MDD-PRS Female N=36; Phase 3, High MDD-PRS Male N=15; Phase 3, Low MDD-PRS Female N=35; Phase 3, Low MDD-PRS Male N=24).

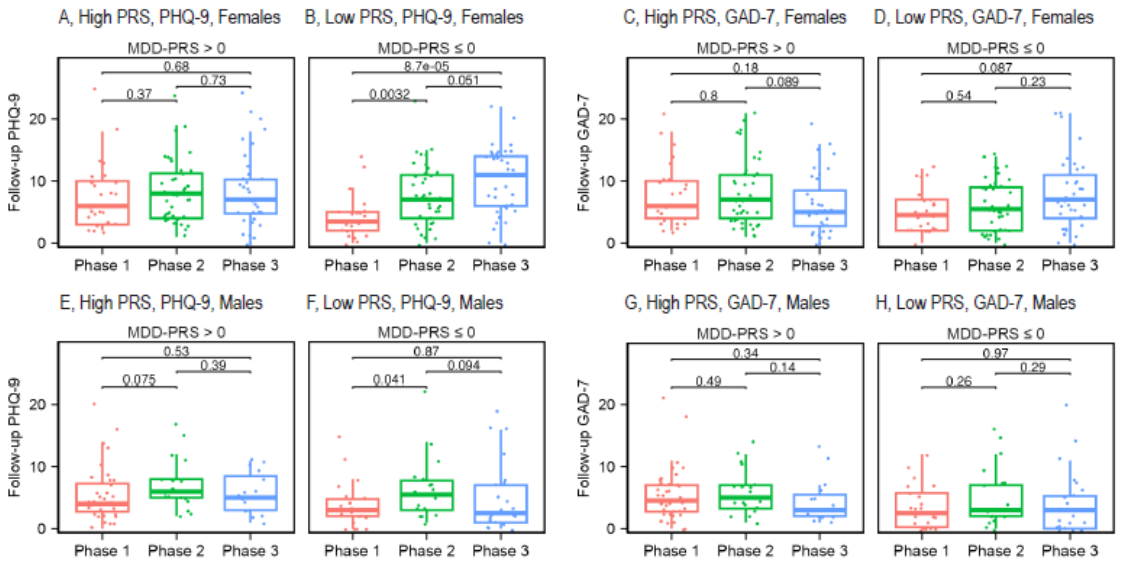

**Supplemental Figure 6. ROC Curve for Logistic Regression Model**

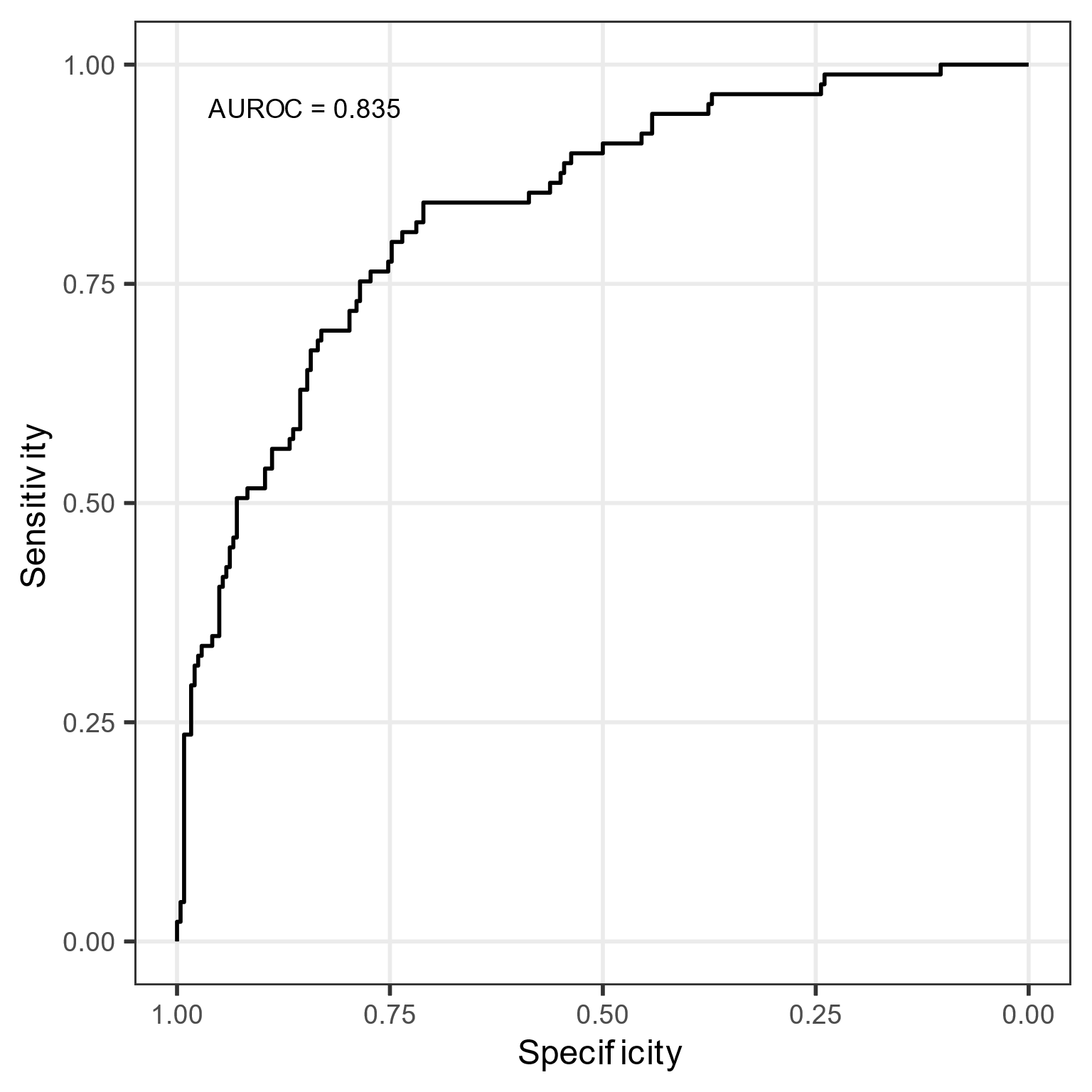

**Supplemental Figure 7. A Multidimensional Scaling Plot of Race for 1000 Genomes and Our Subjects.** All of our subjects (N=331) that self-reported European ancestry were included in the study (OWN-EUR). We did not include any subjects that self-reported other ancestry (OWN-OTHER).

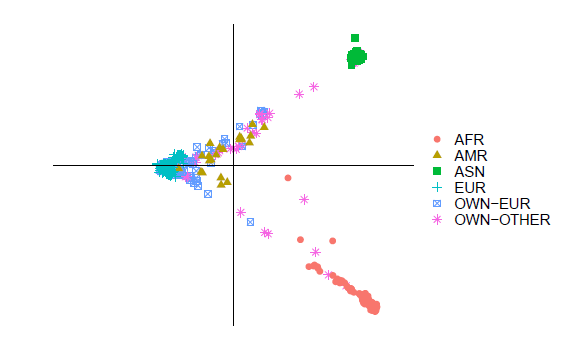

Dimension 2

Dimension 1
